# Early Psychosis Informatics into Care [EPICare]: A co-designed protocol for implementing and evaluating a national integrated digital registry and clinical decision support system within early intervention in psychosis services

**DOI:** 10.1101/2023.06.02.23290899

**Authors:** Siân Lowri Griffiths, Graham K. Murray, Yanakan Logeswaran, John Ainsworth, Sophie M. Allan, Niyah Campbell, Richard Drake, Mohammad Zia Katshu, Matthew Machin, Megan A. Pope, Sarah A. Sullivan, Justin Waring, Tumelo Bogatsu, Julie Kane, Tyler Weetman, Sonia Johnson, James B. Kirkbride, Rachel Upthegrove

**Author notes:** Corresponding Author:* Professor Rachel Upthegrove, Institute for Mental Health, Wolfson Centre, University of Birmingham, 52 Pritchatts Road, Birmingham, B15 2TT. Joint senior authorship (James B. Kirkbride, Rachel Upthegrove). **COMPETING INTERESTS STATEMENT GKM** has received consultancy fees from ieso. RU reports speaker fees from Sunovion, Springer Healthcare, Otsuka and Vitaris outside the submitted work and holds unpaid officership with the British Association for Pharmacology - Honorary General Secretary 2021-2024 and is Deputy Editor, The British Journal of Psychiatry. The views expressed are those of the author(s) and not necessarily those of the NIHR.

## Abstract

**Introduction:** Early Intervention in Psychosis services are nationally mandated in England to provide multidisciplinary care to people experiencing first-episode psychosis, which disproportionately affects deprived and ethnic minority youth. Quality of service provision varies by region, and people from historically underserved populations have unequal access. In other disease areas, including stroke and dementia, national digital registries coupled with clinical decision support systems have revolutionised delivery of equitable, evidence-based interventions to transform patient outcomes and reduce population-level disparities in care and prognosis. Given psychosis is ranked the third most burdensome mental health condition by the World Health Organization, it is essential that we achieve the same parity of health improvements. Here, we provide details of a co-designed protocol to produce an evidence-based, stakeholder-informed framework for the build, implementation, and evaluation of a national integrated digital registry and clinical decision support system for psychosis, known as EPICare (Early Psychosis Informatics for Care).

**Methods and Analysis:** Using a participatory co-design framework, we engaged key stakeholders (N∼40-50) across four meetings to establish the parameters and essential features of EPICare and identify factors likely to influence adoption and implementation into routine practice. Stakeholders consisted of organisational, clinical, academic, and patient and public contributors. In collaboration with National Health Service (NHS) informatics teams, we identified how to retrieve key data items from Electronic Health Records and subsequently design the software architecture and data model to create an infrastructure plan for future implementation. Guided by Normalisation Process Theory, data synthesised from observations of stakeholder meetings and individual interviews (n=10) were subject to interpretative qualitative analysis. Finally, a co-designed set of guides were produced to allow for the build, implementation, and evaluation of EPICare in a larger, future study. An inclusive, representative stakeholder group, fully engaged with the future co-development of EPICare, was also established.

## INTRODUCTION

Psychotic disorders, including schizophrenia, are among the most disabling illnesses worldwide and are often accompanied by enormous personal, family, societal and carer burden (1). Rates of psychosis are unequally distributed throughout the population, with the highest rates found in historically underserved communities, younger populations, and those from minority ethnic backgrounds (2-5). For example, within the UK, people from Black ethnic backgrounds (African, Caribbean, British) are between 3-5 times more likely to experience a first episode of psychosis than White British individuals, and there is evidence that rates are also approximately twice as high for people from Pakistani, Bangladeshi, and mixed ethnic backgrounds in England (2, 6). Further, the need for treatment delivered by Early Intervention Psychosis (EIP) services in England has been identified as highest in several historically underserved regions of England, and in related major conurbations, such as Birmingham, Greater Manchester, Bradford and parts of inner-city London (6). This need for EIP care is closely aligned to populations exposed to greater structural disadvantage including multiple deprivation and social fragmentation (6).

EIP is an internationally adopted model of care, based largely on social inclusion, service user and care engagement, and relapse prevention. In England, EIP services are nationally commissioned to provide evidence-based, multidisciplinary care according to eight NICE-based national standards for people experiencing first-episode psychosis: 1) a maximum waiting time of 14 days from initial referral to commencement of treatment; 2) offer of cognitive behavioural therapy for psychosis; 3) take up of family interventions; 4) offer of clozapine after poor response to at least two other antipsychotic medications; 5) take up of supported employment and education programmes; 6) annual physical health assessments; 7) offer of interventions relevant to physical health (for example, smoking cessation, exercise or substance use programmes); and 8) take up or referral to carer-focused education and support programmes (7). Each care standard is evidence-based, often from randomised controlled trials. Each standard has demonstrated improvement in patient outcomes including remission of symptoms, readmission, recovery, premature mortality and important social and vocational outcomes (8, 9). Importantly, EIP care is cost-effective relative to other forms of care and management for people with psychosis, and EIP services are highly valued by service users (10, 11).

Despite evidence-based standardised targets, only 30-40% of people experiencing psychotic disorders make a full recovery (12), with evidence of large variation in care (13-16). Longer term outcomes are equally poor, with increased rates of physical illnesses (17) and life expectancy reduced by around 15 years compared with people who do not go on to develop severe mental illness (18). This suggests that much work is needed to understand which elements of EIP services are working, and for whom, and whether they lead to better long term outcomes (16).

Variation in outcomes may be related to regional or individual disparities in the care offered and received during EIP, particularly in historically underserved communities where need is greatest, but where there may be insufficient resources to offer standardised care tailored to the needs of local populations. For example, recent data indicates that people with psychosis from Black African and Caribbean backgrounds were 15-30% less likely to receive the equivalent level of cognitive behavioural therapy for their condition compared to White British people (19). Cross-sectional survey data from England and Wales has highlighted further inequalities in care, with Black service users being around 44% less likely to be offered clozapine (19), the only existing medication for treatment-resistant schizophrenia (13). There is also evidence for disparities in outcomes post-EIP, with deprivation related to higher rates of relapse and need for continuing care in secondary mental health services (20). Black and Asian racial minoritized groups are also more likely to continue in secondary mental health care two years following EIP discharge (21).

Despite this, data currently being routinely collected via a patient’s electronic health record does not provide accessible, longitudinal, and nationally representative data to determine the magnitude, causes or consequences of inequitable access to EIP care in England. Relatedly, routine data collected by EIP services in England does not include measures of symptomatic recovery, usually the primary outcome for understanding what treatments work for whom, thus preventing us from developing a national understanding of the clinical effectiveness of treatments in the real world. In turn, neither does it provide a mechanism for immediately improving clinical practice by feeding back real-time actionable insights that would allow treatments to be targeted and tailored to individual patient needs. For example, whilst all EIP providers send data on broad levels of service use into NHS Digital’s Mental Health Services Dataset, the dataset is less suited to ascertain accurate estimates of the incidence of psychotic disorders in England, because current methods of data collection do not differentiate between people engaging in EIP treatment for their first ever episode of psychosis from those who may have existing psychosis, but engage in treatment in a new EIP service for the first time. Further, Mental Health Services Dataset data does not record whether those engaging with EIP treatment later fulfil diagnostic criteria for psychotic disorder. The Mental Health Services Dataset also does not allow us to understand what treatments are delivered to whom, and when, nor their impact on patient recovery and other downstream outcomes.

Furthermore, the pioneering National Clinical Audit of Psychosis (22), which has assessed service fidelity annually since 2017, is a retrospective, cross-sectional manual audit of up to 100 patients with first-episode psychosis in each EIP team in England (22). Although plans exist to revise the data collection methodology, current practice reduces data quality, delays service improvement, and diverts finite EIP resources away from frontline care. There are also no plans for the audit to provide real-time feedback of data to clinical teams. These issues could be eliminated by the provision of a prospectively-collected national digital psychosis registry, able to supply actionable insights in real time to patients, clinical teams, service managers and policymakers via an embedded clinical decision support system (CDSS).

We propose to revolutionise the use of electronic health record data to improve national, local, and individual clinical decision-making and promote better patient and public health outcomes for people experiencing first-episode psychosis, by carefully developing and demonstrating the utility of a prospectively collected digital registry and CDSS in England, capable of being implemented nationally. This would provide standardised information to understand the treated burden of psychosis in the NHS; ensure equitable, responsive, local resource allocation; support reliable, quick, and efficient identification and targeting of any local, regional or group-based disparities in access to care; and improve patient pathways through care and downstream outcomes, including recovery; and finally, enhance understanding of the relationship between interventions provided and outcomes, as well as the relationship between clinical and social characteristics and outcomes.

The potential for further record linkage to other health and social domains also offers the prospect of integrating prospectively collected data from other routine sources including primary care, Office for National Statistics mortality, the Office for National Statistics Census, the National Pupil Database, and Hospital Episode Statistics. This would provide a deeply phenotyped, longitudinal database for clinical and policy decision making. It would also support gold standard research in clinical psychiatry, experimental medicine, and observational epidemiology, to identify, understand, and address the causes and consequences of disparities in health and patient treatment, as well as improve downstream outcomes for people experiencing psychosis.

Digital registries have been deployed successfully in the UK for other disease areas such as stroke, cancer, cystic fibrosis, and dementia (23-28). For example, in the UK, a national stroke registry has transformed patient care and outcomes, with early recognition of different patterns of stroke presentation, focused treatment on previously untreated risk factors, and targeted interventions for improving cognitive impairment (27). In cancer care, tailored interventions based on risk profile have extended lives for thousands of people (28). Yet there are no contemporary examples of digital registries for any secondary care-treated mental health condition listed in the Health Research Classification System mental health category, nor within the international literature, and no specific CDSS for any mental health condition. Integration of a patient-centered digital registry and CDSS for psychosis could be equally transformative and give parity of esteem to one of the most common and disabling set of mental health disorders – psychosis – where there is already a well-developed national infrastructure of EIP services.

To achieve this paradigmatic change in mental health care, our aim is to develop, evaluate and establish a national psychosis registry and CDSS, known as EPICare (Early Psychosis Informatics for Care) in three stages:

- Stage 1: Establish a multidisciplinary and multisector stakeholder network to co-design, de-risk, and define the framework and protocols required to build and implement EPICare as a successful national registry and CDSS.
- Stage 2: Build, pilot, implement, and evaluate the ability of the EPICare platform to improve patient care, enhance service delivery, reduce disparities in care, and demonstrate cost-effectiveness in five demonstrator NHS Trusts, serving underserved and diverse populations with substantial need for EIP care in England.
- Stage 3: Subject to successful implementation and evaluation, seek NHS adoption of EPICare for rollout to all EIP services in England.

### AIMS AND OBJECTIVES

In this paper, we report the protocol for the programme development phase of our activity (Stage 1), in which we aimed to co-design and produce a framework and protocols for onward building, implementation, piloting, and evaluation of a national integrated, patient-centered digital registry and CDSS for psychosis.

To meet this aim, we specifically addressed the following objectives:

1. Establish a network with strong patient and public involvement and engagement (PPIE) and other essential stakeholders to identify essential and desirable elements and minimize unforeseen challenges (Work Package 1).
2. Address key questions on informatics architecture, infrastructure, governance, and integration plans to facilitate onward development and testing of EPICare in diverse NHS Trusts (Work Package 2).
3. Identify implementation factors from the outset to ensure they are considered in designing, implementing, and sustaining the future deployment of EPICare in a measurable way (Work Package 3).

## METHODS AND ANALYSIS

### Study Design

We conducted three concurrent work packages over 12 months, with reciprocal knowledge exchange between work packages, coordinated via fortnightly Programme Management Group meetings. Figure 1 provides a schematic of work packages. The Programme Management Group contained lived experience facilitator, lived experience member, clinicians working in early psychosis and academic members from epidemiology, NHS health informatics, data and implementation science. The EPICare study was reviewed and granted full approval by the Health Research Authority on November 8, 2021 (Ref: 306234).

**Figure 1.**
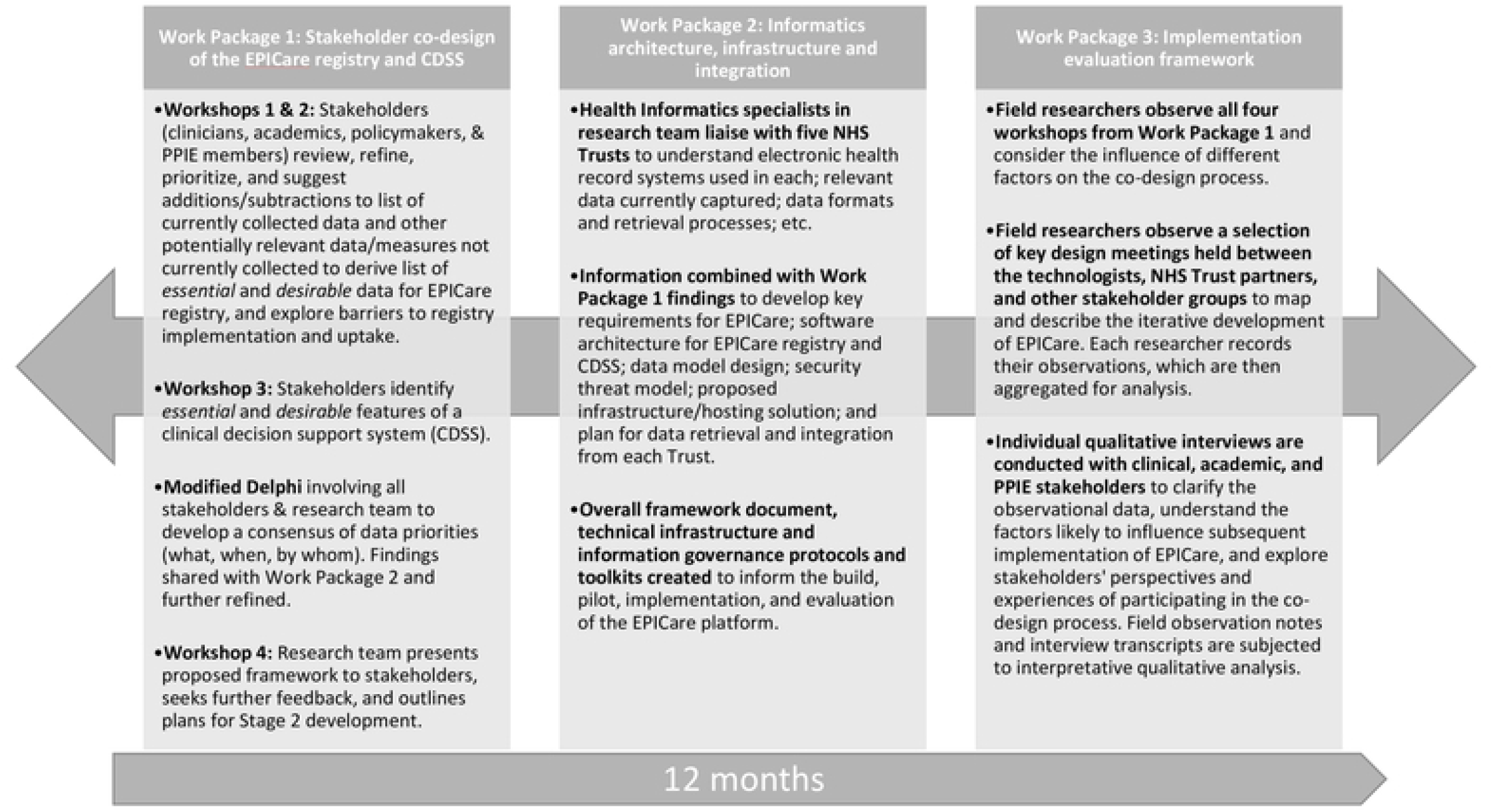
Study design.

#### Work Package 1: Stakeholder co-design of the EPICare registry and CDSS

A participatory co-design framework previously established for informatics in mental health (29) was used to engage a diverse network of stakeholders in the co-design process, including clinicians, academics, policymakers, and PPIE members. Due to the COVID-19 pandemic, workshops were convened online. Whilst this enhanced the scope for collaboration between centres in the study, there were also potential drawbacks of this approach, which included PPIE needing to have access to, and know-how of technology. Second, hosting face-to-face meetings on neutral ground in an approachable format may have helped to removed traditional power structures. The workshops were in a facilitator-led, semi-structured format, including presentations, whole-group discussions, and themed breakout activities (card-sort tasks, small group discussion) with both mixed (random allocation) and streamed group (by broad stakeholder type) sessions on a per-task basis. Essential materials were circulated to stakeholders in advance of each meeting. We also convened additional online preparatory sessions for PPIE stakeholders, led by our PPIE coordinator, to aid understanding and participation in the main workshops. Registry and CDSS goals were examined by stakeholders, who reviewed, refined, and identified a core set of essential and desirable measures that should be collected in the integrated EPICare registry and CDSS, across four domains: sociodemographic measures, treatment measures, patient-reported outcome measures, and clinician-reported outcome measures (CROMS).

To facilitate this process, stakeholders were provided with a list of data already recorded in electronic health records in EIP services, in addition to information on data relevant to the eight nationally mandated NICE standards for EIP care (30) (Health of the Nation Outcomes Scales [HoNOS] on functioning; quality of life and treatment satisfaction [DIALOG], and patient-reported recovery [QPR]). This was supplemented with a minimal set of other initial measures recognized as potentially relevant by the Programme Management Group based on expert knowledge, prior to the first workshop. Examples included symptom ratings, duration of untreated psychosis, and genotyping, amongst others. In Workshops 1 and 2, stakeholders were asked to review, refine, prioritize, and suggest additions or subtractions from this list, with other data that may not currently be routinely collected, but considered by stakeholders to be essential or desirable. The group also explored what barriers to implementation and uptake may be encountered in EPICare (e.g., data security and ownership, time for completion, digitising of routine data currently collected on paper). Similarly, in Workshop 3, stakeholders identified the essential and desirable features of a CDSS to provide timely actionable insights for patients and clinicians, including potential clinician prompts to complete health assessments aligned to NICE standards for EIP care.

After these three initial workshops, we synthesised all information gathered via a modified Delphi approach involving all stakeholders and members of the research team, to develop a consensus of data priorities (what, when, by whom). We shared this with the members of Work Package 2 to understand technical and governance barriers to implementation to further refine our framework to identify a set of “must have” and “could have” data elements. Finally, in Workshop 4, we presented our proposed framework to stakeholders, sought further feedback, and outlined our plans for Stage 2 of EPICare development. From our initial stakeholder network, we sought to retain a representative group of stakeholders for our Stage 2 activity, who will continue to guide the pilot, testing, and evaluation throughout the project, providing a reference point for rapid prototyping and end-user consultation, acceptance, and build.

#### Work Package 2: Informatics architecture, infrastructure and integration: framework, protocol & toolkit

Work Package 2 aimed to reduce technical and governance challenges in the future full build of EPICare by addressing key questions and unknowns. Based on prior experience and knowledge, the area of biggest technical risk for EPICare is the retrieval of data from electronic health records and standardisation of this into a common data model, whilst ensuring compliance with information governance and ethical standards. Previous work by the group involved auditing all EIP services that are part of the National Institute for Health Research (NIHR)-Mental Health Translational Research Collaboration in Early Psychosis (MHTRC-Early Psychosis), to inform understanding of the existing infrastructure, capacity, capabilities, and limitations around designing and developing the EPICare platform for potential national implementation. This initial scoping work has highlighted several different electronic health records in use as well as different ways of capturing and storing relevant data in each of the Trusts.

To build on this knowledge, Health Informatics specialists within the research team contacted and liaised with five NHS Trusts, including Greater Manchester Mental Health NHS Foundation Trust, Birmingham Women’s and Children’s NHS Foundation Trust, Cambridgeshire and Peterborough NHS Foundation Trust, Nottinghamshire Healthcare NHS Foundation Trust and Camden and Islington NHS Foundation Trust. Information Technology team leads and proposed demonstrator sites were identified, gathering information to further understand what electronic health record system is used by each trust; what relevant data are currently captured in the electronic health record (key foci: eight NICE standards for first-episode psychosis treatment); how that data can be retrieved, such as via Application Programming Interface or through regular exports; how the data can, and should be secured during retrieval, complying to the highest information governance standards; and the data formats used for each type of data.

Once this information was captured from all Trusts, it was then used alongside the information gathered from the stakeholders in Work Package 1 to develop and document key requirements for the EPICare system; software architecture for the registry and CDSS; data model design including standardisation of data items into a common format; security threat model including planned treatments for identified threats; proposed infrastructure to include an appropriate hosting solution, such as a secure cloud environment; and an integration plan for retrieval of data from each Trust electronic health record.

This was drafted into an overall framework document and set of technical infrastructure and information governance protocols and toolkits to inform the future build, pilot, implementation, and evaluation of the EPICare platform. With all of this in place, the technical and governance challenges for the main programme grant for applied research application should be significantly reduced.

#### Work Package 3: Implementation evaluation framework

Working in parallel and in collaboration with the members of Work Packages 1 and 2, the purpose of this work package was to establish the preliminary implementation framework for the subsequent testing and roll-out of EPICare. Founded on the idea that implementation research should be integrated throughout all stages of innovation development rather than at ‘end-stage’, this involved understanding the distinct and interconnected implementation issues within the stages of problem definition; iterative evidence-building, intervention conceptualisation, development and testing; and subsequent roll-out, experimentation, and embedding in different service settings. With particular reference to EPICare, this involved understanding how the earlier stages of stakeholder engagement contributed to intervention development and, at the same time, how stakeholders perceived challenges to future adoption and use. With regards to PPIE stakeholders (Work Package 1), this involved understanding views about: a) current challenges in EIP care; b) how clinical registries and CDSS might influence care and service improvement; c) expectations about how interventions might be utilised in standard practice, and d) participants’ experiences of the co-design process. We also studied the early-stage activities of the health informatics team (Work Package 2) to understand the explicit and tacit design assumptions; the contingencies presented by current technological parameters; the influence of prevailing governance arrangements; and importantly, to understand and evidence the interaction between the relative influence of multiple stakeholders in the co-design process. This evidence will be brought together with existing implementation science frameworks, such as Normalisation Process Theory (31), in conjunction with complementary insights drawn from Science and Technology Studies (32, 33).

Normalisation Process Theory helps understand how service innovations are implemented, embedded, and normalised within organisations, to the point where new practices are no longer regarded as new. It is different from other implementation models because it focuses on the specific ‘work’ undertaken by social actors to implement innovations into everyday practice whilst taking into consideration the interplay between actions, contexts, and objects. Normalisation Process Theory has four linked constructs: ‘coherence’, or the work of making sense of an innovation; ‘cognitive participation’, or the work involved when engaging with an innovation; ‘collective action’, or the combined work of integrating new practices into existing skills, relationships, and contexts; and ‘reflexive monitoring’, or the work of continually appraising and adapting to the introduction of new practices. It has been widely used to explain the factors that shape the implementation of complex interventions (31).

Field researchers directly observed all four stakeholder co-design workshops and considered the influence of multiple social, cultural and organisational factors on the co-design process. They also observed a selection of key design meetings held between the technologists, NHS Trust partners, and other stakeholder groups to map and describe the iterative development of EPICare. Each researcher recorded their observations following an agreed semi-structured guide which were then aggregated for analysis.

To clarify the observational data, qualitative semi-structured interviews guided by the constructs from Normalisation Process Theory were then conducted with all stakeholder groups to understand the factors likely to influence subsequent implementation of EPICare. An initial set of questions and topics derived from the study objectives were used to systematically code interview transcripts and develop themes. This was piloted on four transcripts by two researchers, before agreeing to a revised set of codes, followed by further coding of remaining transcripts. Interviews focused on the different cognitive-cultural perspectives of each stakeholder group, their experiences of participating in the co-design process, and their perceptions about their influence on the co-design, together with their recommendations for subsequent development and testing.

#### Study Participants

Participants were recruited between November and December 2021. We recruited forty participants across all stakeholder workshops (Work Package 1). This included at least ten people with lived experience of psychosis, and ideally, lived experience of early intervention, to form the PPIE stakeholder group. PPIE members were recruited from the Birmingham University Youth Advisory Group, Cambridgeshire & Peterborough Foundation Trust, Bristol Lived Experience Advisory Panel and PPIE networks at University College London, including those associated with the National Institute for Health Research Mental Health Policy Research Unit. As an acknowledgement of the time and effort involved in taking part in the study, PPIE participants were reimbursed in line with the INVOLVE payment policy (www.involve.org.uk), which equates to £25 per hour of participation.

The remaining thirty participants were recruited from the breadth of multidisciplinary care in EIP services (psychiatrists, psychologists, occupational therapists, social workers, and nurses), in addition to stakeholders from the charitable sector, NHS England, policymakers, and other academics, for facilitated group meetings. The clinical collaborators were recruited from NHS Trusts serving diverse and underserved areas with a combined population of approximately 3.73m people (10.1% of the English population at risk for psychosis): Birmingham Women’s and Children’s Trust, Manchester Health and Care NHS Foundation Trust, Camden and Islington NHS Foundation Trust, Avon and Wiltshire Mental Health Partnership Trust, and Cambridgeshire & Peterborough Foundation Trust. Attendance at the stakeholder group meetings was taken as consent for this process and no individual written consent was required from stakeholders (including PPIE).

All stakeholders were also invited to participate in individual qualitative interviews in Work Package 3 to ensure that we selected a representative subset of each stakeholder group from our Work Package 1 stakeholder meetings. Written informed consent was obtained and interviewees were given a unique participant identification number, which was used throughout the transcription of interviews to ensure anonymity.

Given the online group format of the stakeholder workshops, individual participants attending these workshops were identifiable to each other and to the authors. However, the identities of participants who consented to an individual qualitative interview were known only to the interviewer, and as noted above, interviewees were assigned a unique participant identification number to ensure their anonymity during the transcription of their interviews.

#### Analysis

All interviews were recorded and transcribed. For quality control, transcript summaries were shared with participants and feedback elicited as to their veracity. Observation notes of the stakeholder workshops and transcripts of the individual interviews were subject to interpretative qualitative analysis, guided by the Normalisation Process Theory implementation science framework.

Preliminary data analysis of observation notes involved producing short descriptive summaries of field observations, for the purpose of summarising and sharing data with the study team. NVivo software was used to organise and analyse the qualitative observational and transcribed interview data. An iterative coding process was followed with data being subject to systematic close reading and coding. Through sharing and deliberating preliminary codes and interpretations with the wider study team and through the processes of constant comparison, secondary inductive and interpretative themes were developed. At this stage, the constructs of Normalisation Process Theory were used to further analyse and explain the study findings. Through discussion and disputation with PPIE, clinicians, and the project team, inferences were made about how the implementation science framework should be further refined.

## DISCUSSION

The purpose of this study is to develop, implement and evaluate a national integrated, patient-centered digital registry and CDSS for psychosis (EPICare) to improve national, local and individual clinical decision-making and promote improved outcomes for people experiencing first-episode psychosis. The EPICare registry and CDSS potentially represent a paradigmatic shift, as they would be the first patient-centered digital registry and CDSS for psychosis, one of the most common and disabling mental health disorders disproportionately affecting deprived and disadvantaged youth. By combining routine, systematic, prospective data collection via a national digital registry with real-time actionable insights delivered to patients, clinical teams, service managers and policymakers via an embedded CDSS, EPICare aims to improve patient care, enhance service delivery, reduce disparities in care, and further our understanding of the relationship between the interventions offered to, and received by, young people receiving EIP care and outcomes. A further aim of the study is to demonstrate cost-effectiveness in five demonstrator NHS Trusts serving underserved and diverse populations with substantial need for EIP care in England.

In this paper, we have reported the protocol for the programme development phase of this study (Stage 1), in which we aimed to co-design and produce a framework and protocols for onward building, implementation, piloting, and evaluation of the EPICare registry and CDSS. Strengths of this first phase of the study include the use of a participatory design to co-design a framework for EPICare with input from diverse relevant stakeholders, including lived experience experts and clinical, academic, technologist and organisational stakeholders. By engaging multiple stakeholders in an iterative co-design process and using qualitative methods to capture and synthesise rich data representing a variety of perspectives, we have succeeded in establishing a network with strong patient and public involvement and engagement (PPIE) and representation from other essential stakeholder groups and in collaboratively identifying essential and desirable elements of the EPICare platform (Work Package 1). We have also worked to proactively identify and minimize potential challenges and barriers to uptake and implementation (Work Package 3), including by addressing key questions related to informatics architecture, infrastructure, governance, and integration in diverse NHS Trusts (Work Package 2).

While we have achieved all of the objectives set out for the first phase of this study, it is worth noting that adoption and integration of all the desirable platform elements identified by stakeholders may not be feasible or pragmatic for the initial build of the data model. This will be tested in our next stage.

Next steps for the EPICare study include Stage 2 building, piloting, implementation, and evaluation of the EPICare platform in five demonstrator NHS Trusts serving underserved and diverse populations with substantial need for EIP care in England. If successful, this will be followed by Stage 3, in which we will seek NHS adoption of EPICare for rollout to all EIP services in England.

## Data Availability

No datasets were generated or analysed during the current study. All relevant data from this study will be made available upon study completion.

## ETHICS AND DISSEMINATION

The EPICare programme has been granted full approval by the Health Research Authority (Ref: 306234).

The results will be disseminated through peer-reviewed journals, conference presentations, media outlets, the internet, and various community / stakeholder engagement activities.

## AUTHORS’ CONTRIBUTIONS

R.U. and J.B.K. are joint project leads and hold joint senior authorship. S.L.G. drafted the manuscript with further input from R.U. and J.B.K., S.J., G.K.M., M.A.P., N.C., J.W., J.A., R.D., M.M., S.A., T.J.W., J.K., Y.L., T.B. and S.S. also contributed to the study as well as provided comments on the manuscript. All authors approved the final version.

